# Estimating the Economic Burden of Typhoid Fever in Ibadan, Nigeria

**DOI:** 10.64898/2026.11.12.26367962

**Authors:** Yunjin Yum, Janet Ogundairo, Folashayo P. Adeniji, Gabriel Nyirenda, Pius Ifiok Udofia, Seung Eun Kyung, Veronica Olatimbo Ogunleye, Mohamadou Siribie, Ondari Mogeni, Oluwafemi Popoola, Hyonjin Jeon, Birkneh Tilahun Tadesse, Iruka N. Okeke, Florian Marks, Jung-Seok Lee, Aderemi Kehinde

## Abstract

**Background:** Evidence on the economic burden of typhoid fever remains limited in sub-Saharan Africa. This study estimated the costs associated with typhoid fever and related invasive *Salmonella* infections in Ibadan, Nigeria.

**Methods:** A field-based cost-of-illness study was conducted at six healthcare facilities between 2022 and 2025. Patients with *Salmonella enterica* serovar Typhi (*S*. Typhi), *Salmonella enterica* serovar Paratyphi (*S*. Paratyphi), invasive non-typhoidal *Salmonella* (iNTS), or typhoid gastrointestinal perforation (special cases) were enrolled, with malaria patients serving as controls. Direct medical, direct non-medical, and indirect costs were estimated for each episode and converted from Nigerian naira to US dollars using year-specific exchange rates for 2022–2025. Cases and controls were matched by age group, sex, and point of care, and confidence intervals were estimated using non-parametric bootstrapping.

**Results:** Among 198 participants, 69 were *S*. Typhi cases, one was *S*. Paratyphi case, 14 were iNTS cases, 12 were special cases, and 102 were malaria controls. Average total costs per episode were $130 for *S*. Typhi, $27 for *S*. Paratyphi, $156 for iNTS, $583 for special (perforation) cases, and $52 for malaria. For *S*. Typhi, costs were substantially higher among inpatients than outpatients ($309 vs. $72). Compared with matched malaria controls, the mean incremental total costs were $42 (95% CI: 18–71) for *S*. Typhi, $104 (95% CI: 25–197) for iNTS, and $560 (95% CI: 289–882) for special (perforation) cases.

**Conclusions:** Typhoid fever imposes a considerable economic burden in Ibadan, Nigeria, particularly among patients requiring hospitalization or experiencing severe complications. These findings provide locally relevant evidence for evaluating typhoid conjugate vaccine introduction and informing resource-allocation decisions.

## Introduction

Typhoid fever is a systemic bacterial infection caused by *Salmonella enterica* serovar Typhi (*S*. Typhi), transmitted primarily through contaminated food and water under conditions of poor sanitation and hygiene (1, 2). Although the disease has been virtually eliminated in high-income countries with safe water and sanitation infrastructure, it remains a persistent public health challenge across low- and middle-income countries (LMICs), particularly in Asia and sub-Saharan Africa (3). Global estimates suggest approximately 9 million cases and 110,000 deaths occur each year, with the highest burden concentrated in regions where access to safe water and effective waste management is limited (4). Beyond its health consequences, typhoid fever imposes substantial economic costs on patients, households, healthcare providers, and society through medical expenditures and productivity losses (5) (6).

To address this continuing threat, typhoid conjugate vaccines (TCV) have been developed and shown to be safe and highly effective (7). In 2018, the World Health Organization (WHO) recommended introducing TCV in countries with endemic or drug-resistant *S*. Typhi (8). Following the WHO recommendation, 12 countries have introduced TCV into their national immunization programs: Pakistan (2019), Liberia (2021), Zimbabwe (2021), Samoa (2021), Nepal (2022), Malawi (2023), Fiji (2023), Tuvalu (2024), Burkina Faso (2025), Kenya (2025), Niger (2025), and Bangladesh (2025). Sierra Leone and India are planning national introduction (9).

Despite these advances, many endemic countries without improved water and sanitation have not yet introduced TCV (10). A challenge in prioritizing typhoid prevention strategies is the limited availability of context specific economic evidence to inform policymakers about the value of vaccination and Water Sanitation and Hygiene (WASH) relative to other pressing health priorities (6, 11). Without robust economic assessments, decision-makers lack the data needed to understand the potential returns on investment and to set rational policy priorities.

Evidence on the economic burden of typhoid fever is especially sparse in Africa. A recent review of published cost-of-illness (COI) studies highlighted that most analyses to date have been conducted in Asian countries, while evidence from African settings remains limited, with only a few ongoing or unpublished studies expected to fill this gap (5). The absence of standardized, field-based COI data limits the comparability of findings across regions and hinders regional and global policy discussions (12). Therefore, generating such evidence in African countries is critical.

Nigeria represents one of the most affected countries in sub-Saharan Africa. A systematic review reported that the prevalence of typhoid fever varies widely, from 2% to 50%, depending on the region and diagnostic approach. Poor access to potable water, inadequate waste disposal, and widespread inappropriate antibiotic use contribute to sustained transmission and the emergence of multidrug-resistant *S*. Typhi strains (13). Typhoid fever is endemic in Ibadan, Nigeria, where this study was conducted, 2017 to 2019 surveillance recorded an incidence of 36 per 100,000 person years (14-16). Typhoid burdens are likely higher elsewhere in Nigeria but difficult to estimate given poor access to diagnostics and care (17). The rise of resistant strains worldwide, and including in Nigeria (18) (19), is associated with more severe outcomes, including bloodstream infections, prolonged hospitalizations, and increased mortality, further straining already resource-limited health systems.

Estimating the COI of typhoid fever in Nigeria is therefore essential. COI studies can capture direct medical costs, direct non-medical costs, and productivity losses, providing insights into the financial impact of typhoid fever from patient and health system perspectives. These data are critical for cost-effectiveness analyses of TCV introduction and for informing resource allocation in a context where health budgets are constrained and competing priorities are many. To address this gap, we conducted a COI study with the primary objective of estimating the economic burden of typhoid fever in Ibadan, Nigeria. Secondary analyses described the costs associated with *Salmonella enterica* serovar Paratyphi (*S*. Paratyphi), invasive non-typhoidal *Salmonella* (iNTS), and gastrointestinal perforation cases, while patients with malaria served as controls. The study aimed to strengthen the existing evidence base and inform policy discussions on preventive measures, including vaccination, to reduce the burden of typhoid fever in Nigeria and similar settings.

## Methods

### Study design

The study was a prospective hospital-based COI study conducted in Ibadan, Oyo State, Nigeria, at six health facilities, four of which had previously participated in the Severe Typhoid in Africa (SETA) surveillance program (Table 1) (20). The six health facilities included in the study were as follows: University College Hospital (UCH), located in northern metropolitan Ibadan, is the country’s first teaching hospital and a major referral center with approximately 1,445 beds. Our Lady of Apostles (OLA) Catholic Hospital in Oluyoro is the largest private hospital in the city, with about 160 beds, providing medical, surgical, pediatric, obstetric, and gynecologic care. Adeoyo Maternity Teaching Hospital, Yemetu is a long-standing public facility that delivers maternity, pediatric, gynecologic, and general medical services through multiple wards and specialized units, employing over 500 staff. Additional sites included the Kola Daisi Foundation (KDF) Primary Healthcare Centre in Ibadan North with 19 beds, which provides pediatrics, obstetrics, antenatal care, immunization, and family planning services; the Idi Ogungun Primary Health Care Centre in Ibadan South East with 16 beds, which offers outpatient consultations, maternal and child health care, and disease surveillance; and the Oranyan Primary Health Care Centre, also in Ibadan South East, which provides laboratory diagnostics, child welfare, family planning, antenatal and immunization clinics, and human immunodeficiency virus and tuberculosis services supported by 49 medical staff, including permanent staff, ad hoc workers, and volunteers.

**Table 1.** Study areas, study period and number of cases.

| **Health facility for study** | **Study period** | **No. of cases** |
|---|---|---|
| University College Hospital Ibadan | Oct, 2022 - Nov, 2025 | 51 |
| Our Lady of Apostles Catholic Hospital | May, 2023 - Dec, 2025 | 15 |
| Adeoyo Maternity Teaching Hospital | Aug, 2023 - Sep, 2025 | 20 |
| Kola Daisi Foundation Primary Healthcare Centre | Oct, 2022 - Jul, 2025 | 34 |
| Idi Ogungun Primary Health Care | Nov, 2023 - Nov, 2025 | 33 |
| Oranyan Primary Health Care | Feb, 2024 - Nov, 2025 | 45 |

### Participants and eligibility

Participants were classified into five case-type categories: *S*. Typhi, *S*. Paratyphi, iNTS, special (perforation) cases, and malaria for controls. *S*. Typhi, *S*. Paratyphi, and iNTS cases were defined based on blood culture-confirmed infection with the corresponding pathogen. Special (perforation) cases included participants with clinically diagnosed typhoid gastrointestinal perforation for which no other cause of perforation was confirmed, even in the absence of laboratory confirmation. Malaria cases were defined as participants whose blood cultures were negative for bacterial pathogens or contaminants but who had laboratory-confirmed malaria based on a positive rapid diagnostic test (RDT) or blood smear microscopy.

At the time of the study, neither malaria vaccines (which are now being phased in Nigeria) nor typhoid conjugate vaccines (which have not been adopted) were in use. Antimicrobial sensitivity testing was conducted using Kirby-Bauer disc diffusion method according to Clinical and Laboratory Standards Institute guidelines (14) and strains resistant to Ampicillin, Azithromycin, Chloramphenicol, Ciprofloxacin, Ceftriaxone, and Cotrimoxazole were classified as resistant for the purpose of the analysis.

### Cost-of-Illness Estimation

The analysis incorporated two components: (i) patient out-of-pocket (OOP) expenditures and productivity losses, assessed through patient-level COI surveys (private cost), and (ii) public costs, representing the health facility perspective, were defined as treatment costs not covered by patients’ OOP payments and estimated through a review of hospital billing records.

COI interviews were conducted with patients meeting the inclusion criteria up to four times throughout the course of their illness. Interviews were scheduled relative to the date of enrollment (Day 0), when participants visited the study facility and provided blood samples for diagnosis. The initial interview was conducted on Days 3–7, followed by interviews on Days 12–14, 28–30, and 90 ± 7. At the time of the first interview, participants were interviewed regarding all direct medical and non-medical out-of-pocket expenditures, income losses, substitute laborer costs, and caretaker costs incurred up to the day before the interview, and socioeconomic information such as household income and expenditure was also collected. If the patient continued to feel ill after the first interview, a second interview was conducted to collect subsequent OOP expenditures and indirect costs incurred between the first interview and the day before the second interview. To minimize recall bias, participants were provided with a diary card to record treatment-related costs, additional healthcare visits, productivity losses, and support from substitute laborers or caregivers between interviews. A third interview was conducted two weeks later (approximately Day 28–30), if the patient still felt sick.

Although a fourth follow-up interview was originally planned for Day 90 for participants with prolonged illness, no participants required a fourth interview at this stage. At the end, all cost data were consolidated to estimate the total cost per episode by case types. Relapse episodes occurring within 90 days of a previous blood culture-confirmed typhoid fever, paratyphoid fever, or iNTS were treated as a continuation of the initial illness and their costs were combined. Episodes occurring 90 days or more after a previous blood culture-confirmed infection were classified as re-infections, considered new illness episodes, and assigned separate study identifiers.

The current study captured three main cost components: direct medical costs (DMC), direct non-medical costs (DNMC), and indirect costs (IC). DMC covered all expenses directly related to medical care for the illness episode, including consultation, medications, diagnostics, and other facility-based services. OOP payments were collected through patient interviews. Non-OOP payments (public costs) were extracted from hospital invoices as the portion of the total bill not paid directly by the patient.

These costs included amounts financed through insurance schemes and government subsidies, as well as project-funded laboratory services. By capturing payments made by both private and public payers, the analysis aimed to estimate the full range of costs associated with each illness episode and their distribution between private and public sectors.

DNMC comprised non-treatment expenses such as transportation, food, and accommodation incurred by patients and their companions. IC included productivity loss of the patients, caregiver and substitute labor costs. Human capital approach was used to estimate productivity losses associated with illness. This approach values time lost from productive activities based on forgone earnings and the opportunity cost of unpaid work (21, 22). For patients, caregivers, and substitute laborers engaged in paid work, productivity losses were estimated using their reported daily wages. For those engaged in unpaid work, opportunity costs were valued using the daily minimum wage in Nigeria (NGN 1,500 until 2023, and 3,500 until 2025) (23). For students, opportunity costs were valued using their reported daily tuition fees. The number of days applied to these valuations reflected the degree of activity loss: each day of complete inability to perform usual activities was counted as a full day, whereas each day of partial inability was counted as half day.

Where available, patient-reported medical expenditures were cross-checked against hospital invoices. Data quality was further supported by ongoing monitoring and regular discussions with data collectors throughout the study period.

### Statistical analysis

Continuous variables were summarized using means and standard deviations (SD), whereas categorical variables were presented as frequencies and percentages. Missing public cost values (approximately 13%) were imputed using the mean public cost among participants with the same case type and point of care. For participants with missing monthly household income data (approximately 25%), household asset ownership was compared with the typical asset profiles of the three income groups defined by tertiles among participants with reported income. These participants were assigned to the most comparable income group and imputed with the mean income of that group.

To compare costs between *Salmonella*-related diagnostic group (*S*. Typhi, iNTS, and special (perforation) cases) and the malaria group within the same demographic and care-setting strata, one-to-one exact matching was performed based on age group (0–4, 5–9, 10–14, 15–19, and ≥20 years), gender and point of care (inpatient versus outpatient). Age group and gender were used to improve demographic comparability, while point of care was included to account for substantial cost differences between inpatient and outpatient care. The matched analysis was intended as a conditional cost comparison rather than a causal effect analysis. Mean cost differences between matched cases and controls were estimated.

Given the typically skewed distribution of cost data, 95% confidence intervals (CIs) for cost estimates were derived using a non-parametric bootstrap resampling approach with 1,000 iterations and the percentile method (2.5th and 97.5th percentiles) (24, 25). All costs were converted to USD using official World Bank exchange rates for the corresponding study years (426 per USD in 2022, 645 per USD in 2023, and 1,479 per USD in 2024 and 2025) (26). All statistical analyses were conducted using Stata version 18.0.

### Ethics statement

Ethical approval for the study was received from the Institutional Review Boards of the International Vaccine Institute (IVI) (Approval number 2020-009), the University of Ibadan/ University College Hospital Ethics Committee (Approval Number UI/EC/21/0190), Oyo State Health Research Ethics Committee (Approval Number NHREC/OYOSHRIEC/10/11/12) and OLACH Ethics Committee (Approval Number OC/EC/21/10). Patients enrolled in the COI studies, provided written informed consent. For participants younger than 18 years, written consent was obtained from a parent or caretakers, and assent was additionally obtained from participants aged 12–17 years.

## Results

Table 2 shows the baseline characteristics of patients by case type. The mean age was below 12 years across all case types. The proportion of inpatients was highest among special (perforation) cases, followed by iNTS, *S*. Typhi, and malaria. The mean number of sick days was highest among special (perforation) cases, followed by *S*. Typhi, iNTS, *S*. Paratyphi, and malaria. Although patients with *S*. Typhi had more total sick days than those with iNTS, the proportion of full days lost relative to total sick days was higher among patients with iNTS. No patients reported employing substitute labor during their illness, whereas the majority across all five case types had caregivers. The average monthly household income varied across case types, ranging from $115 among *S*. Paratyphi to $274 among special (perforation) cases. Median values (minimum– maximum) for the continuous variables are presented in Table S1.

**Table 2.** Baseline characteristics of participants.

|  | ***S*. Typhi (N=69)** | ***S*. Paratyphi (N=1)** | **iNTS (N=14)** | **Special (perforation) cases (N=12)** | **Malaria (N=102)** |
|---|---|---|---|---|---|
| Age, y | 8.9 (6.5) | 1.0 (–) | 7.8 (18.6) | 11.8 (3.1) | 8.9 (6.9) |
| Male | 38 (55.1%) | 1 (100.0%) | 7 (50.0%) | 4 (33.3%) | 49 (48.0%) |
| Inpatient | 17 (24.6%) | 0 (0.0%) | 4 (28.6%) | 12 (100.0%) | 10 (9.8%) |
| Number of sick days prior to enrollment | 7.1 (5.2) | 2.0 (–) | 4.4 (3.2) | 7.9 (3.9) | 3.5 (2.9) |
| Total sick days | 15.6 (6.7) | 6.0 (–) | 11.5 (9.3) | 20.8 (8.1) | 5.8 (4.0) |
| Number of full days lost due to illness | 7.1 (7.3) | 0.0 (–) | 6.4 (9.2) | 12.2 (6.0) | 1.6 (2.3) |
| Number of partial days lost due to illness | 6.8 (5.6) | 6.0 (–) | 3.3 (2.6) | 8.2 (7.8) | 3.6 (4.0) |
| Patients with wage loss | 2 (2.9%) | 0 (0.0%) | 0 (0.0%) | 0 (0.0%) | 3 (2.9%) |
| Patients with carers | 65 (94.2%) | 1 (100.0%) | 14 (100.0%) | 12 (100.0%) | 98 (96.1%) |
| Patients with substitute labor cost for carers | 0.0 (0.0) | 0 (0.0%) | 0.0 (0.0) | 0.0 (0.0) | 0.0 (0.0) |
| Monthly household income (US\$) | 204 (277) | 115 (–) | 223 (258) | 274 (321) | 240 (283) |

Note. Continuous variables are presented as mean (SD), and binary variables are presented as n (%).

Figure 1 illustrates the distribution of patient costs by case type and the share of private versus public payments. For most of the case types, DMC represented the largest proportion of total OOP expenditure, followed by IC (Figure 1(A)). For *S*. Paratyphi cases, IC costs were not reported and DNMC accounted for the next largest share. In Figure 1(B), private direct medical costs accounted for the majority of DMC in all case types. The difference between private and public contributions was smallest for malaria, where the two sources contributed nearly equally.

**Figure 1.**
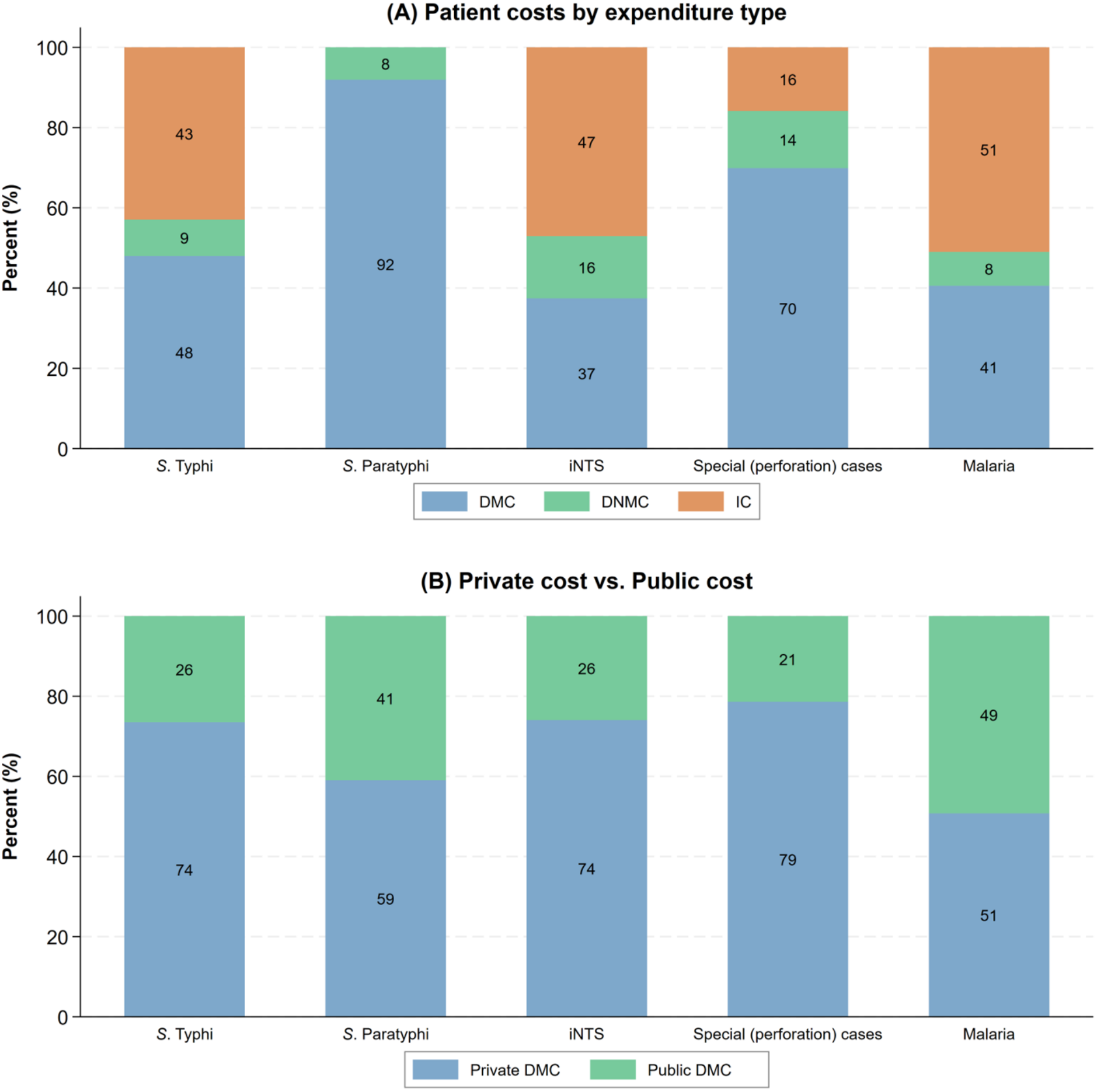
Proportion of the economic burden for type of cases by (A) expenditure type and (B) payer type.

Table 3 summarizes the average economic burden per episode. The total cost per outpatient (OPD) episode was $72 for *S*. Typhi, $27 for *S*. Paratyphi, $98 for iNTS, $40 for malaria; no outpatient special (perforation) cases were observed. In the case of inpatient (IPD), the highest total cost per episode was special (perforation) cases ($583), followed by *S*. Typhi ($309), iNTS ($302), and malaria ($163). Direct medical, direct non-medical, and indirect costs generally followed the same pattern, with special (perforation) cases incurring the highest costs and malaria the lowest. In particular, DMC was notably high in special (perforation) cases, likely because patients diagnosed with pathognomonic gastrointestinal perforations in this group were more likely to have undergone surgical procedures. Across all disease categories, inpatient episodes had higher costs than outpatient episodes. When costs were standardized per day, costs for OPD episodes ranged from $5 (*S*. Typhi and *S*. Paratyphi) to $11 (iNTS) and IPD episodes ranged from $17 (malaria) to $31 (special (perforation) cases). Median values (minimum–maximum) for the economic burden per episode are presented in Table S2 and total costs by case type based on WGS classification are presented in Table S3.

**Table 3.** Average economic burden per episode (US$).

|  | ***S*. Typhi** |  | ***S*. Paratyphi** |  | **iNTS** |  | **Special (perforation) cases** |  | **Malaria** |  |
|---|---|---|---|---|---|---|---|---|---|---|
|  | **OPD (N=52)** | **IPD (N=17)** | **OPD (N=1)** | **IPD (N=0)** | **OPD (N=10)** | **IPD (N=4)** | **OPD (N=0)** | **IPD (N=12)** | **OPD (N=92)** | **IPD (N=10)** |
|  | Mean (CI<sup>b</sup>: lower, upper) |  | Mean (CI: lower, upper) |  | Mean (CI: lower, upper) |  | Mean (CI: lower, upper) |  | Mean (CI: lower, upper) |  |
| Direct Medical Cost (DMC)<sup>a</sup> | 33 (28–41) | 194 (141–255) | 26 (–)<sup>c</sup> | – | 37 (29–45) | 152 (107–201) | – | 436 (312–587) | 23 (21–25) | 93 (64–131) |
| Direct Non-Medical Cost (DNMC) | 4 (3–7) | 27 (24–31) | 1 (–) | – | 4 (1–8) | 66 (22–117) | – | 70 (50–94) | 2 (1–2) | 18 (8–32) |
| Indirect Cost (IC) | 35 (26–45) | 88 (39–153) | 0 (–) | – | 57 (3–161) | 84 (21–147) | – | 78 (50–113) | 16 (12–20) | 53 (22–96) |
| Total Cost | 72 (60–85) | 309 (227–391) | 27 (–) | – | 98 (38–208) | 302 (192–396) | – | 583 (450–739) | 40 (36–46) | 163 (100–258) |
| Total Cost per Day<sup>d</sup> | 5 (4–6) | 18 (13–24) | 5 (–) | – | 11 (5–19) | 21 (14–28) | – | 31 (22–41) | 10 (8–11) | 17 (11–26) |

<sup>a</sup> DMC reflects the combined amount of private and public expenditure
<sup>b</sup> Bootstrapping with the percentile method
<sup>c</sup> The 95% confidence interval was not estimated for *S*. Paratyphi because only one patient was included.
<sup>d</sup> Total cost divided by total number of sick days

**Table 4.** Mean differences in economic burden between matched cases and controls (US$).

|  | ***S*. Typhi (n=59) vs. Malaria (n=59)** | **iNTS (n=13) vs. Malaria (n=13)** | **Special (perforation) cases (n=5) vs. Malaria (n=5)** |
|---|---|---|---|
|  | Mean (CI<sup>b</sup>: lower, upper) | Mean (CI: lower, upper) | Mean (CI: lower, upper) |
| Direct Medical Cost (DMC)<sup>a</sup> | 18 (7–31) | 34 (12–59) | 454 (188–796) |
| Direct Non-Medical Cost (DNMC) | 3 (1–6) | 18 (2–43) | 62 (47–77) |
| Indirect Cost (IC) | 21 (4–42) | 51 (-9–138) | 44 (-25–117) |
| Total Cost | 42 (18–71) | 104 (25–197) | 560 (289–882) |

<sup>a</sup> DMC reflects the combined amount of private and public expenditure
<sup>b</sup> Bootstrapping with the percentile method

Following one-to-one exact matching on age group, gender, and point of care, 59, 13, and 5 matched pairs were identified for *S*. Typhi, iNTS, and special (perforation) cases, respectively. *S*. Paratyphi was excluded from the matched case–control analysis because only one case was enrolled. The mean differences in total cost between each case type and the matched controls were $42 (95% CI: 18–71), $104 (95% CI: 25–197), and $560 (95% CI: 289–882), respectively. As the bootstrap-derived 95% confidence intervals for total cost excluded zero for all case types, total costs were higher among cases than matched controls.

To examine whether the relative economic burden of *Salmonella*-related cases varied by household income level, private expenditure was estimated as a ratio of monthly household income and are presented in by income tertile in Figure 2. Across all expenditure categories, the economic burden was highest among low-income households, where DMC, DNMC, and IC accounted for 367%, 57%, and 61% of monthly household income, respectively. The corresponding proportions were lower, but still considerable, among middle-income (53%, 12%, and 29%) and high-income households (21%, 6%, and 22%).

**Figure 2.**
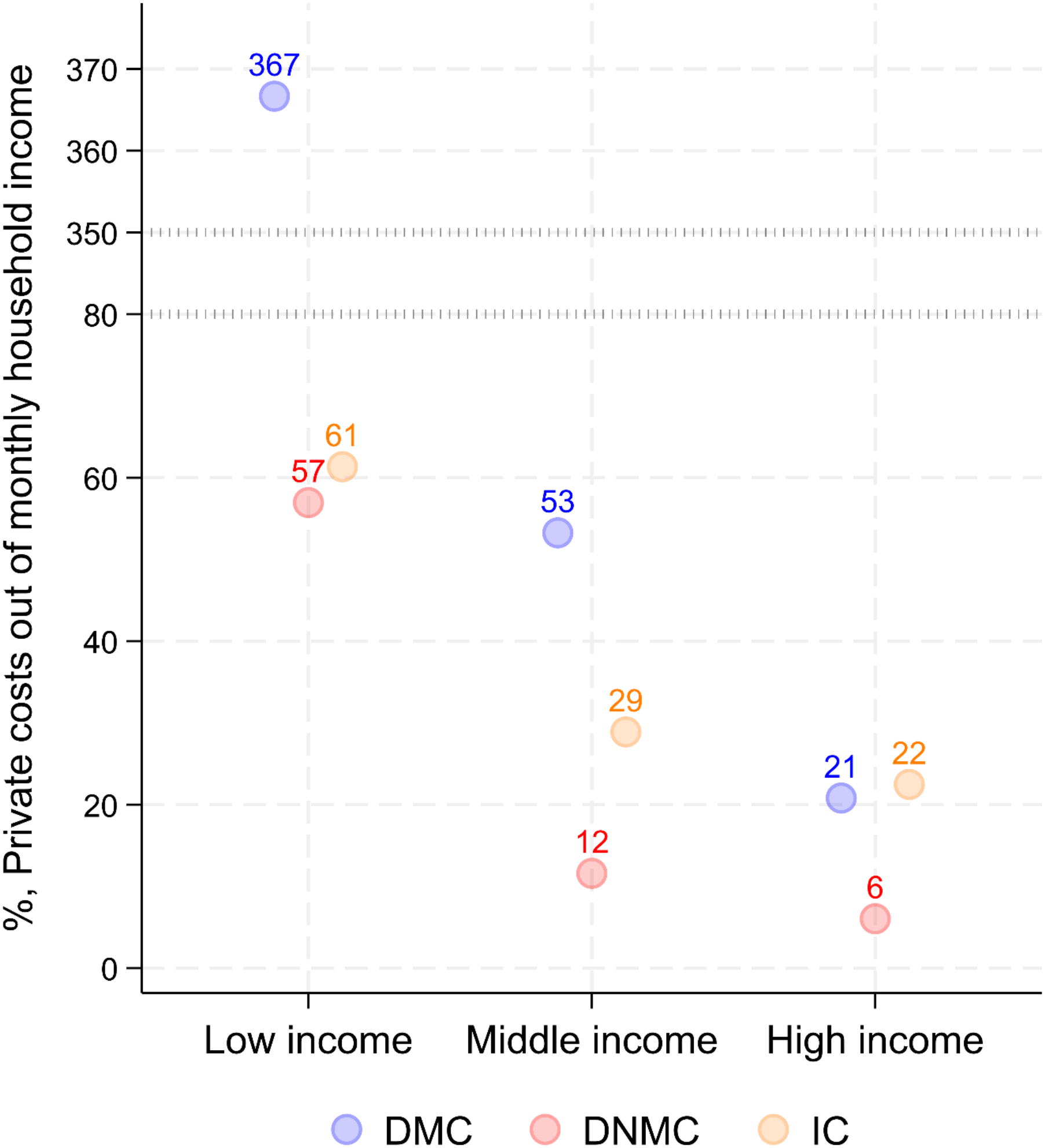
Ratio of economic burden of Salmonella-related cases (S. Typhi, S. Paratyphi, iNTS, and special (perforation) cases) to monthly household income (n=96).

Figure 3 compares the average economic burden of susceptible and resistant *S*. Typhi and iNTS. Resistant *S*. Typhi and iNTS cases tended to have higher costs than susceptible cases. The average total cost per episode was $72 for susceptible *S*. Typhi and $135 for resistant *S*. Typhi, and $48 for susceptible iNTS and $174 for resistant iNTS. However, these findings should be interpreted with caution, particularly for susceptible cases (*S*. Typhi n=5; iNTS n=2), due to the small sample size.

**Figure 3.**
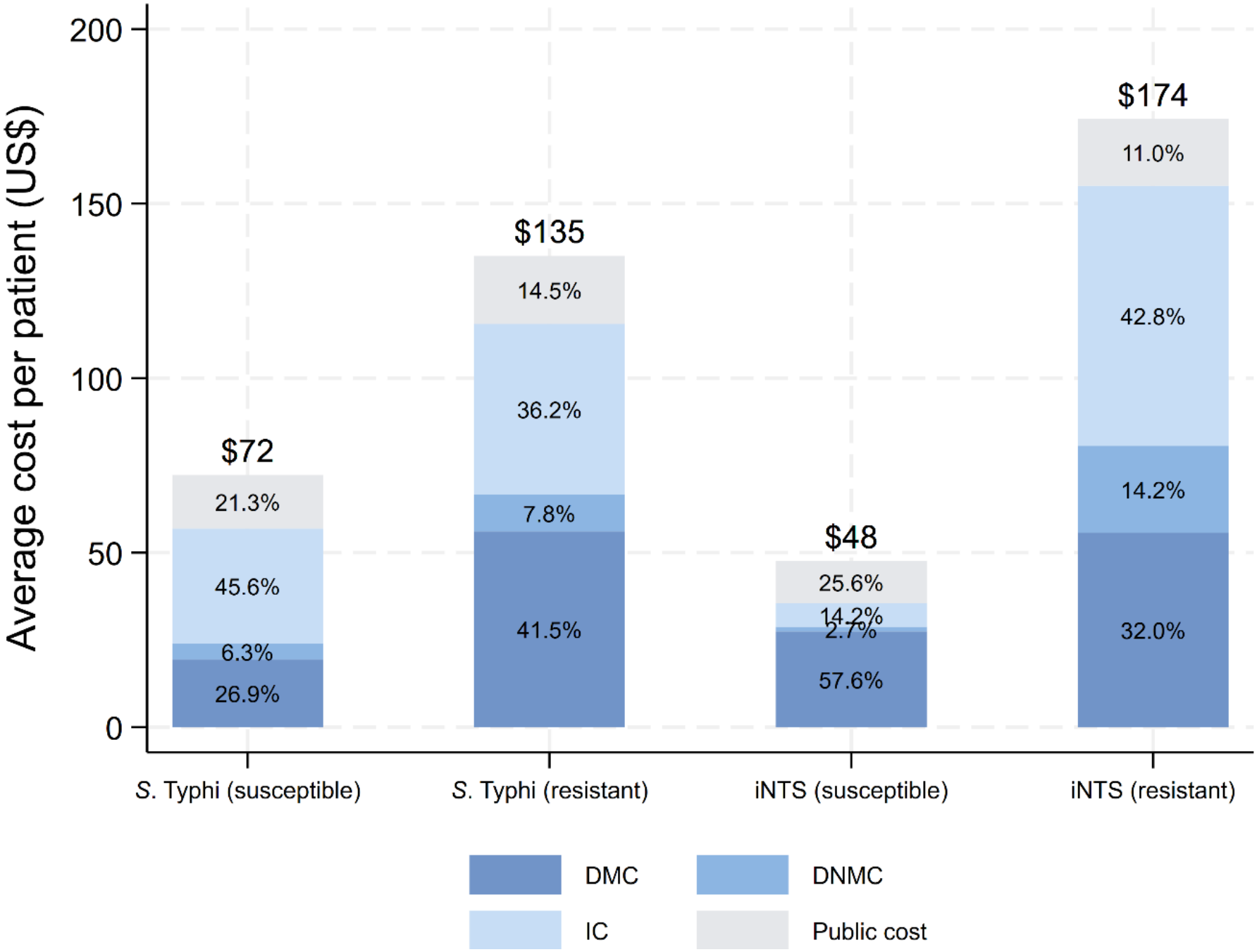
Average economic burden per episode among susceptible and resistant S. Typhi and iNTS cases. DMC reflects the amount of private expenditure; Sample sizes were as follows: susceptible S. Typhi (n=5), resistant S. Typhi (n=64), susceptible iNTS (n=2), and resistant iNTS (n=12).

## Discussion

This study provides empirical estimates of the economic burden of typhoid fever in Ibadan, Nigeria using patient-level, field-based COI data that integrate private direct medical, direct non-medical, indirect, and public costs. Across all case types, patients bore the majority of costs directly through out-of-pocket payments, underscoring the substantial economic burden and the need to enhance insurance coverage in Nigeria. Only 5% of the patients in this study had public or civil insurance coverage and there is a pressing need to move the National Health Insurance Authority (covering 22 million of the estimated 200 million Nigerians at time of writing) towards its stated goal of Universal Health Coverage.

The total cost per episode varied widely by disease severity, ranging from $27 for *S*. Paratyphi to nearly $583 for special (perforation) cases. All patients classified as special (perforation) cases were inpatients and underwent surgical procedures, reflecting the higher severity of illness. Overall, the economic burden tended to increase with disease severity. Furthermore, the matched case-control analysis showed that typhoid cases incurred substantially higher costs than matched malaria controls, even after adjusting for age, gender, and point of care.

Direct medical costs constituted the largest component of the economic burden. The burden of illness was particularly pronounced among low-income households, where illness-related costs accounted for substantially greater share of household income than among middle- or high-income households. These findings suggest that typhoid fever can impose considerable financial hardship on affected households, particularly among economically vulnerable populations. The burden may be especially severe when hospitalization is required, given the substantially higher costs associated with inpatient episodes.

We observed that the number of days to presentation (enrollment) was longer for patients from whom *S*. Typhi was cultured, than for those yielding iNTS. We speculate that this arises from the fact that the iNTS patients are mostly at extremes of age (median age 4 years, with 6/ <2 years and one >70 years), while *S*. Typhi patients were aged 18 months to 34, median 8 years. This higher median among febrile patients from whom *S*. Typhi was cultures approaches that of the special (perforation) cases, who were also mostly late presenters. The data suggest that late presentation may contribute to severity and cost, and that any intervention that prevents Typhi infection and perforation in children above 2 years could eliminate these burdens.

Several studies have previously examined the economic burden of typhoid fever across Asia and Africa. In South and Southeast Asia, multiple published studies have reported the economic burden of typhoid fever (6, 27-29). Among them, one study that used patient surveys and hospital records as data sources reported inpatient cost was $159 for India and $531 for Indonesia. The estimates observed in the present study were generally within the range reported in the literature, although household income is substantially smaller in Ibadan, compared to those locations. However, direct comparisons should be interpreted cautiously because of differences in study design, healthcare systems, and costing methodologies. For example, in that study, different costing approaches were applied across study sites, including the use of expert opinion to estimate treatment costs. In Africa, a study conducted in Tanzania reported an estimated cost of $171 per episode (including both inpatients and outpatients), which was comparable to the findings of the present study (30). In Malawi, mean household costs were $94 for inpatients and $19 for outpatients, while healthcare provider costs were higher at $297 and $40, respectively. Unlike in our study, household costs in Malawi were lower than healthcare provider costs, potentially reflecting the provision of free healthcare through government facilities (31).

The findings of the present study have several important policy implications. First, they provide essential inputs for evaluating the cost-effectiveness of TCV introduction in Nigeria. The high cost per episode, particularly for severe and complicated cases, suggests that reductions in incidence through vaccination could generate cost savings for households and the health system (32). Second, the study contributes to understanding the broader economic implications of antimicrobial resistance (AMR) in typhoid. In the present study, resistant cases tended to incur higher costs than susceptible cases, with average total costs of $135 versus $72 for *S*. Typhi and $174 versus $48 for iNTS, respectively, although these findings should be interpreted with caution given the limited number of susceptible cases. The findings suggest that the rise of antibiotic-resistant *S*. Typhi may contribute to greater treatment complexity and higher costs, potentially through the need for more expensive or prolonged antibiotic regimens. This may increase the societal burden, both directly (through higher medical expenses) and indirectly (through longer illness and hospitalization). In this context, introducing TCVs, as well as improving WASH, could have broader economic benefits by preventing infections that would otherwise necessitate costly antimicrobial treatment, thereby supporting national AMR containment strategies. Additionally, the cost of illness from unprevented typhoid will continue to grow as resistance expands more broadly.

Several limitations should be acknowledged. First, the study was limited to six health facilities in Ibadan, which is not representative of all regions of Nigeria particularly rural areas with different healthcare-seeking patterns and cost structures. Outside the South West geopolitical zone, in which Ibadan lies, access to care is greatly reduced and economic costs are likely to be substantially greater (17). In addition, the numbers of S. Paratyphi, iNTS, and special (perforation) cases were relatively small, and therefore cost estimates for these groups should be interpreted with caution. Second, despite the use of diary cards, recall bias cannot be entirely excluded, especially for expenditures incurred outside the study sites. To minimize this bias, regular data verification and quality checks were conducted in collaboration with site teams throughout the study period to enhance data completeness and reliability. Third, some cost and socioeconomic data were missing because of limited access to facility recording systems or participants’ reluctance to disclose financial information. In such cases, missing values were imputed using available information. Nevertheless, these approaches may have introduced some degree of measurement error.

Despite these limitations, the current study provides robust, locally relevant evidence on the economic impact of typhoid fever in Nigeria. It contributes to the evidence needed to guide vaccine introduction and resource allocation. Future research should extend this work to other regions and incorporate prospective follow-up to capture long-term economic and social consequences of the disease.

## Conclusion

Typhoid fever imposed substantial costs in Ibadan, particularly among hospitalized patients and those with gastrointestinal perforation, and costs were higher than among malaria controls matched on age group, sex, and point of care. Patient costs were largest relative to monthly household income among low-income households, highlighting the potential for considerable financial strain. These locally derived estimates can inform evaluations of TCV introduction and other typhoid-control strategies, including WASH interventions, although studies in other regions of Nigeria are needed to assess generalizability.

## Data Availability

Data supporting this article are available from the corresponding author upon reasonable request.

## Acknowledgments

The authors would like to thank Lilian Nsere Danbagai, Olubusayo Odewale and Adekemi Adepeju for technical support and helpful contributions.

## References

1. Mudau M, Ngobeni-Nyambi R, Momba MNB. The Fascinating Cross-Paths of Pathogenic Bacteria, Human and Animal Faecal Sources in Water-Stressed Communities of Vhembe District, South Africa. Pathogens. 2023;12(9).

2. Musa DA, Dim-Gbereva L, Ogbiko C, Nwodo OF. Phytochemical and in vitro anti-typhoid properties of leaf, stem and root extracts of Ficus capensis (Moraceae). Journal of Pharmacy & Bioresources. 2019;16(2):165–72.

3. Cutler D, Miller G. The role of public health improvements in health advances: the twentieth-century United States. Demography. 2005;42(1):1–22.

4. Institute for Health Metrics and Evaluation. Global Burden of Disease (GBD): Global Burden of Disease; 2023 [Available from: https://vizhub.healthdata.org/gbd-results/].

5. Debellut F, Friedrich A, Baral R, Pecenka C, Mugisha E, Neuzil KM. The cost of typhoid illness in low- and middle-income countries, a scoping review of the literature. PLOS ONE. 2024;19(6):e0305692.

6. Luthra K, Watts E, Debellut F, Pecenka C, Bar-Zeev N, Constenla D. A Review of the Economic Evidence of Typhoid Fever and Typhoid Vaccines. Clinical Infectious Diseases. 2019;68(Supplement_2):S83–S95.

7. Shakya M, Colin-Jones R, Theiss-Nyland K, Voysey M, Pant D, Smith N, et al. Phase 3 Efficacy Analysis of a Typhoid Conjugate Vaccine Trial in Nepal. N Engl J Med. 2019;381(23):2209–18.

8. World Health O. Typhoid vaccines: WHO position paper, March 2018 - Recommendations. Vaccine. 2019;37(2):214–6.

9. International Vaccine Access Center (IVAC), Johns Hopkins Bloomberg School of Public Health, International Vaccine Access Center (IVAC). VIEW-hub [Available from: www.view-hub.org].

10. Hancuh M. Typhoid fever surveillance, incidence estimates, and progress toward typhoid conjugate vaccine introduction—worldwide, 2018–2022. MMWR Morbidity and mortality weekly report. 2023;72.

11. Watson CH, Edmunds WJ. A review of typhoid fever transmission dynamic models and economic evaluations of vaccination. Vaccine. 2015;33:C42–C54.

12. Lee J-S, Mogasale V, Lim JK, Ly S, Lee KS, Sorn S, et al. A multi-country study of the economic burden of dengue fever based on patient-specific field surveys in Burkina Faso, Kenya, and Cambodia. PLOS Neglected Tropical Diseases. 2019;13(2):e0007164.

13. Akinyemi KO, Oyefolu AOB, Mutiu WB, Iwalokun BA, Ayeni ES, Ajose SO, et al. Typhoid Fever: Tracking the Trend in Nigeria. Am J Trop Med Hyg. 2018;99(3_Suppl):41–7.

14. Marks F, Im J, Park SE, Pak GD, Jeon HJ, Nana LRW, et al. Incidence of typhoid fever in Burkina Faso, Democratic Republic of the Congo, Ethiopia, Ghana, Madagascar, and Nigeria (the Severe Typhoid in Africa programme): a population-based study. The Lancet Global Health. 2024;12(4):e599–e610.

15. Sunmonu GT, Ogunleye VO, Ikhimiukor OO, Popoola OA, Osadebamwen PE, Kehinde A, et al. Slow-growing Salmonella enterica serovariety Typhi mis-identified as serovariety Gallinarum in Ibadan, Nigeria. PLOS Glob Public Health. 2025;5(7):e0004745.

16. Kehinde PA, Ogunleye DV, Popoola DO, Amadi ME, Coker MA, Rahman MM, et al. Seasonal Patterns in Typhoid Environmental Surveillance in Ibadan, Nigeria. International Journal of Infectious Diseases. 2025;152:107386.

17. Abubakar I, Dalglish SL, Angell B, Sanuade O, Abimbola S, Adamu AL, et al. The Lancet Nigeria Commission: investing in health and the future of the nation. The Lancet. 2022;399(10330):1155–200.

18. Carey ME, Dyson ZA, Ingle DJ, Amir A, Aworh MK, Chattaway MA, et al. Global diversity and antimicrobial resistance of typhoid fever pathogens: Insights from a meta-analysis of 13,000 Salmonella Typhi genomes. Elife. 2023;12:e85867.

19. Ikhimiukor OO, Oaikhena AO, Afolayan AO, Fadeyi A, Kehinde A, Ogunleye VO, et al. Genomic characterization of invasive typhoidal and non-typhoidal Salmonella in southwestern Nigeria. PLOS Neglected Tropical Diseases. 2022;16(8):e0010716.

20. Park SE, Toy T, Cruz Espinoza LM, Panzner U, Mogeni OD, Im J, et al. The Severe Typhoid Fever in Africa Program: Study Design and Methodology to Assess Disease Severity, Host Immunity, and Carriage Associated With Invasive Salmonellosis. Clinical Infectious Diseases. 2019;69(Supplement_6):S422–S34.

21. Rice DP. Estimating the cost of illness. American Journal of Public Health and the Nations Health. 1967;57(3):424–40.

22. Pike J, Grosse SD. Friction Cost Estimates of Productivity Costs in Cost-of-Illness Studies in Comparison with Human Capital Estimates: A Review: J. Pike, SD Grosse. Applied health economics and health policy. 2018;16(6):765–78.

23. Federal Ministry of Labour and Employment. Clarification of the 2019 National Minimum Wage Bill 2019 [Available from: https://labour.gov.ng/2019/02/05/clarification-of-the-2019-national-minimum-wage-bill/].

24. Lee JS, Carabali M, Lim JK, Herrera VM, Park IY, Villar L, et al. Early warning signal for dengue outbreaks and identification of high risk areas for dengue fever in Colombia using climate and non-climate datasets. BMC Infect Dis. 2017;17(1):480.

25. Nixon RM, Wonderling D, Grieve RD. Non-parametric methods for cost-effectiveness analysis: the central limit theorem and the bootstrap compared. Health Econ. 2010;19(3):316–33.

26. Official exchange rate (LCU per US$, period average) 2026 [Available from: https://data.worldbank.org/indicator/PA.NUS.FCRF?locations=NG].

27. Kaljee LM, Pach A, Garrett D, Bajracharya D, Karki K, Khan I. Social and Economic Burden Associated With Typhoid Fever in Kathmandu and Surrounding Areas: A Qualitative Study. J Infect Dis. 2018;218(suppl_4):S243–s9.

28. Cook J, Jeuland M, Whittington D, Poulos C, Clemens J, Sur D, et al. The cost-effectiveness of typhoid Vi vaccination programs: calculations for four urban sites in four Asian countries. Vaccine. 2008;26(50):6305–16.

29. Poulos C, Riewpaiboon A, Stewart JF, Clemens J, Guh S, Agtini M, et al. Cost of illness due to typhoid fever in five Asian countries. Tropical medicine & international health. 2011;16(3):314–23.

30. Riewpaiboon A, Piatti M, Ley B, Deen J, Thriemer K, Von Seidlein L, et al. Cost of illness due to typhoid fever in Pemba, Zanzibar, East Africa. Journal of Health, Population, and Nutrition. 2014;32(3):377.

31. Limani F, Smith C, Wachepa R, Chafuwa H, Meiring J, Noah P, et al. Estimating the economic burden of typhoid in children and adults in Blantyre, Malawi: A costing cohort study. PLOS ONE. 2022;17(11):e0277419.

32. Ryckman T, Karthikeyan AS, Kumar D, Cao Y, Kang G, Goldhaber-Fiebert JD, et al. Comparison of Strategies for Typhoid Conjugate Vaccine Introduction in India: A Cost-Effectiveness Modeling Study. J Infect Dis. 2021;224(Supple 5):S612–s24.

