## Supplementary materials for "Estimating the Economic Burden of Typhoid Fever in Ibadan, Nigeria"

*Table S1. Baseline characteristics presented as medians (min-max).*

| | *S.* Typhi (N=69) | *S.* Paratyphi (N=1) | iNTS (N=14) | Special (perforation) cases (N=12) | Malaria (N=102) |
|---|---|---|---|---|---|
| Age, y | 8 (2-34) | 1 (-) | 2 (0-72) | 12 (7-16) | 8 (0-41) |
| Number of sick days prior to enrollment | 5 (0-29) | 2 (-) | 4 (1-14) | 7 (3-14) | 3 (0-24) |
| Total sick days | 14 (5-39) | 6 (-) | 10 (4-41) | 18 (14-42) | 5 (1-31) |
| Number of full days lost due to illness | 5 (0-32) | 0 (-) | 2 (0-34) | 12 (2-22) | 0 (0-10) |
| Number of partial days lost due to illness | 5 (0-28) | 6 (-) | 4 (0-8) | 6 (1-27) | 3 (0-26) |
| Monthly household income (US$) | 123 (10-1596) | 115 (-) | 135 (14-883) | 137 (31-1089) | 123 (8-1705) |

*Table S2. Economic burden per episode presented in medians (min-max).*

| | *S.* Typhi | | *S.* Paratyphi | | iNTS | | Special (perforation) cases | | Malaria | |
|---|---|---|---|---|---|---|---|---|---|---|
| | OPD (N=52) | IPD (N=17) | OPD (N=1) | IPD (N=0) | OPD (N=10) | IPD (N=4) | OPD (N=0) | IPD (N=12) | OPD (N=92) | IPD (N=10) |
| | Median (Min – Max) | | Median (Min – Max) | | Median (Min – Max) | | Median (Min – Max) | | Median (Min – Max) | |
| Direct Medical Cost (DMC)[a] | 25 (11-174) | 183 (22-521) | 26 (-)[c] | – | 35 (18-61) | 146 (89-227) | – | 336 (227-1196) | 21 (5-52) | 85 (13-220) |
| Direct Non-Medical Cost (DNMC) | 3 (0-46) | 28 (7-42) | 1 (-) | – | 1 (1-19) | 50 (17-146) | – | 59 (22-175) | 1 (0-7) | 13 (3-64) |
| Indirect Cost (IC) | 22 (0-158) | 40 (9-473) | 0 (-) | – | 5 (0-517) | 82 (19-155) | – | 69 (9-196) | 9 (0-112) | 23 (6-200) |
| Total Cost | 63 (18-261) | 282 (80-708) | 27 (-) | – | 41 (26-578) | 343 (125-397) | – | 523 (271-1316) | 34 (7-139) | 142 (22-484) |

| | | | | | | | | | | |
|---|---|---|---|---|---|---|---|---|---|---|
| Total Cost per Day[b] | 5 (1-17) | 12 (6-40) | 5 (-) | – | 6 (2-44) | 22 (10-29) | – | 28 (9-66) | 7 (1-59) | 12 (4-44) |

[a] DMC reflects the combined amount of private and public expenditure

[b] Total cost divided by total number of sick days

[c] Min-Max was not estimated for *S*. Paratyphi because only one patient was included.

*Table S3. Average economic burden per case type based on WGS.*

| **Case type based on WGS** | **N** | **Mean (SD)** | **Median (Min - Max)** |
|---|---|---|---|
| *S*.Typhi | 69 | 130 (141) | 80 (18 - 708) |
| *S*.Paratyphi | 1 | 27 (-) | - |
| iNTS | 14 | 156 (181) | 54 (26 - 578) |
| Special (perforation) cases | 12 | 583 (284) | 523 (271 - 1316) |
| Malaria | 102 | 52 (58) | 38 (7 - 484) |
| Total | 198 | 119 (176) | 50 (7 - 1316) |

WGS; Whole Genome Sequencing, SD; standard deviation.
